# Standardization and Comparison of Emergency Use Authorized COVID-19 Assays and Testing Laboratories

**DOI:** 10.1101/2023.11.08.23297633

**Authors:** Anuradha Rao, Jessica Lin, Richard Parsons, Morgan Greenleaf, Adrianna Westbrook, Eric Lai, Heather B. Bowers, Kaleb McClendon, William O’Sick, Tyler Baugh, Markayla Sifford, Julie A. Sullivan, Wilbur A. Lam, Leda Bassit

**Affiliations:** The Atlanta Center for Microsystems-Engineered Point-of-Care Technologies, Atlanta, GA, United States of America; Department of Pediatrics, Emory University School of Medicine, Atlanta, Georgia, USA; Wallace H. Coulter Department of Biomedical Engineering, Emory University and Georgia Institute of Technology, Atlanta, GA, USA; Nell Hodgson Woodruff School of Nursing, Emory University, Atlanta, GA USA; Emory University School of Medicine, Atlanta, GA, USA; Personalized Science San Diego CA 05403 USA; Laboratory of Biochemical Pharmacology, Emory University, Atlanta, Georgia; Emory/Children’s Laboratory for Innovative Assay Development, Atlanta, Georgia, USA; Department of Pathology and Laboratory Medicine, Emory University School of Medicine Atlanta, GA USA; Children’s Healthcare of Atlanta, Atlanta, Georgia, USA; Aflac Cancer and Blood Disorders Center at Children’s Healthcare of Atlanta, Atlanta, Georgia, USA

**Keywords:** Standardization, comparator, COVID-19, SARS-CoV-2, testing, calibration, molecular, antigen

## Abstract

SARS-CoV-2, the causal agent of the global COVID-19 pandemic, made its appearance at the end of 2019 and is still circulating in the population. The pandemic led to an urgent need for fast, reliable, and widely available testing. After December 2020, the emergence of new variants of SARS-CoV-2 led to additional challenges since new and existing tests had to detect variants to the same extent as the original Wuhan strain. When an antigen-based test is submitted to the Food and Drug Administration (FDA) for Emergency Use Authorization (EUA) consideration it is benchmarked against PCR comparator assays, which yield cycle threshold (C_T_) data as an indirect indicator of viral load – the lower the C_T_, the higher the viral load of the sample and the higher the C_T_, the lower the viral load. The FDA mandates that 10-20% of clinical samples used to evaluate the antigen test have to be low positive. Low positive, as defined by the FDA, are clinical samples in which the C_T_ of the SARS-CoV-2 target gene is within 3 C_T_ of the mean C_T_ value of the approved comparator test’s Limit of Detection (LOD). While all comparator tests are PCR-based, the results from different PCR assays used are not uniform. Results vary depending on assay platform, target gene, LOD and laboratory methodology. The emergence and dominance of the Omicron variant further challenged this approach as the fraction of low positive clinical samples dramatically increased as compared to earlier SARS-CoV-2 variants. This led to 20-40% of clinical samples having high C_T_ values and therefore assays vying for an EUA were failing to achieve the 80% Percent Positive Agreement (PPA) threshold required. Here we describe the methods and statistical analyses used to establish a predefined cutoff, based on genome copies per ml (GE/ml) to classify samples as low positive (less than the cutoff GE/ml) or high positive (greater than the cutoff GE/mL). C_T_ 30 for the E gene target using Cobas® SARS-CoV-2-FluA/B platform performed at TriCore Reference Laboratories, and this low positive cutoff value was used for two EUA authorizations. Using droplet digital PCR and methods described here, a value 49,447.72 was determined as the GE/ml equivalent for the low positive cutoff. The C_T_ cutoff corresponding to 49447.72 GE/ml was determined across other platforms and laboratories. The methodology and statistical analysis described here can now be used for standardization of all comparators used for FDA submissions with a goal towards establishing uniform criteria for EUA authorization.

**Motivation:** The motivation for this work was the need to establish a predefined cutoff based on genome copies per ml (GE/ml) rather than Ct, which can vary depending on the laboratory and assay used. A GE/ml-based threshold was necessary to define what constituted ‘low positives” for samples that were included in data sets submitted to the FDA for emergency use approval for SARS-CoV-2 antigen tests.

## Introduction

Coronavirus Disease 2019 (COVID-19) is a severe febrile respiratory illness caused by Severe Acute Respiratory Syndrome Coronavirus 2 (SARS-CoV-2). The first documented cases of COVID-19 were reported in Wuhan City, China, in December 2019.^1^ The first molecular diagnostic tests for SARS-CoV-2 were developed shortly afterwards, and utilized reverse-transcription polymerase chain reaction (RT-PCR) for highly sensitive detection of viral RNA.^2^ The first rapid-antigen tests (RATs) to detect viral protein would not be developed until months later.^3,4^ Due to their high sensitivity, RT-PCR tests remain the gold standard for COVID-19 diagnosis.

To facilitate the development of diagnostic tests, the Food and Drug Administration (FDA) issued an Emergency Use Authorization (EUA) in May 2020. As a part of EUA submissions for antigen tests, the FDA requires tests to be validated using clinical samples, and further require 10-20% of clinical samples used to validate the tests to be low positive. Low positives are defined by the FDA as samples in which a gene target is within 3 cycle thresholds (C_T_s) of the mean C_T_ value of the Limit of Detection (LoD) of a RT-PCR-based comparator test.^3,5,6^ For example, if the mean LOD C_T_ of the comparator is 33.0, then 10-20% of the clinical samples used for EUA submission must have a Ct of 30 or higher. However, these requirements presented several issues for test developers.

First, despite its status as the gold standard for COVID-19 diagnosis, RT-PCR is a semi-quantitative measure, and C_T_ values vary based on choice of assay and platform, and differences in laboratory methods^7,8^. Thus, despite the high sensitivity of comparator tests, there is still some variation in the Ct values, which can affect the definition of ‘low positive’ for the evaluation new tests.

Secondly, it has been shown that viral antigens and RNA have different kinetics over the course of infection, and antigen tests are less sensitive to samples with low Ct values.^9–12^ This further complicates the relationship between antigen- and nucleic-acid-based tests, as the correlation between the two may change based on the point in time the sample is taken and patient characteristics such as age, symptomaticity, and immunization status.^13–15^ Thus, there is a need for a standard reference not subject the variation of clinical patient samples.

A third issue arose during the latest Omicron wave of COVID-19. The arrival of the Omicron variant, and the increased immunity among the population led to an increase in C_T_ > value of patient samples, which correlates to a decrease in viral load.^16,17^ Data suggest that the percentage of samples with C_T_ > 30 increased from December 2021 to January 2022 with the appearance of the Omicron variant. Prevalence of low positive samples were observed at increased frequency in several clinical studies conducted during early in 2022 (20 – 40%). Namely, it appeared that the Omicron variant increased the measured C_T_ value in the comparator RT-PCR assays due to a decrease in the viral load per sample.^12,16,18–22^ This shift in Ct values further muddies the term “low positive,” emphasizing the need for a more quantitative standard when evaluating tests.

To adapt to a potentially changing virus, the FDA provided a “controlled analysis” approach for PPA calculations in EUA antigen assay submissions by using a pre-defined C_T_ value to classify COVID-19 samples into high positives (C_T_ < 30) or low positives (C_T_ > 30). This cutoff was used for two antigen rapid LFA tests granted EUA, Osang and Xiamen Boson tests.^23,24^ A standardized measurement of viral genome equivalents per milliliter (GE/ml) is needed to allow a controlled analysis of several assays and platforms to serve as comparators for antigen test evaluations. Other methods to harmonize the C_T_ values stop at standardizing Ct value for better agreement between laboratories and tests, without further extrapolating to regulatory standards, and in particular the regulatory standards of antigen tests.^25–28^

In this resource, we establish a method for generating calibration curves to establish the relationship between GE/mL and C_T_ across multiple assays, with the goal of standardizing comparators across assays, platforms, and testing laboratories for better antigen test evaluation (**Figure 1**). In addition, we set a quantitative cut-off in GE/mL for “low positive” samples that has been used in regulatory approvals by the FDA.

**Figure 1.**
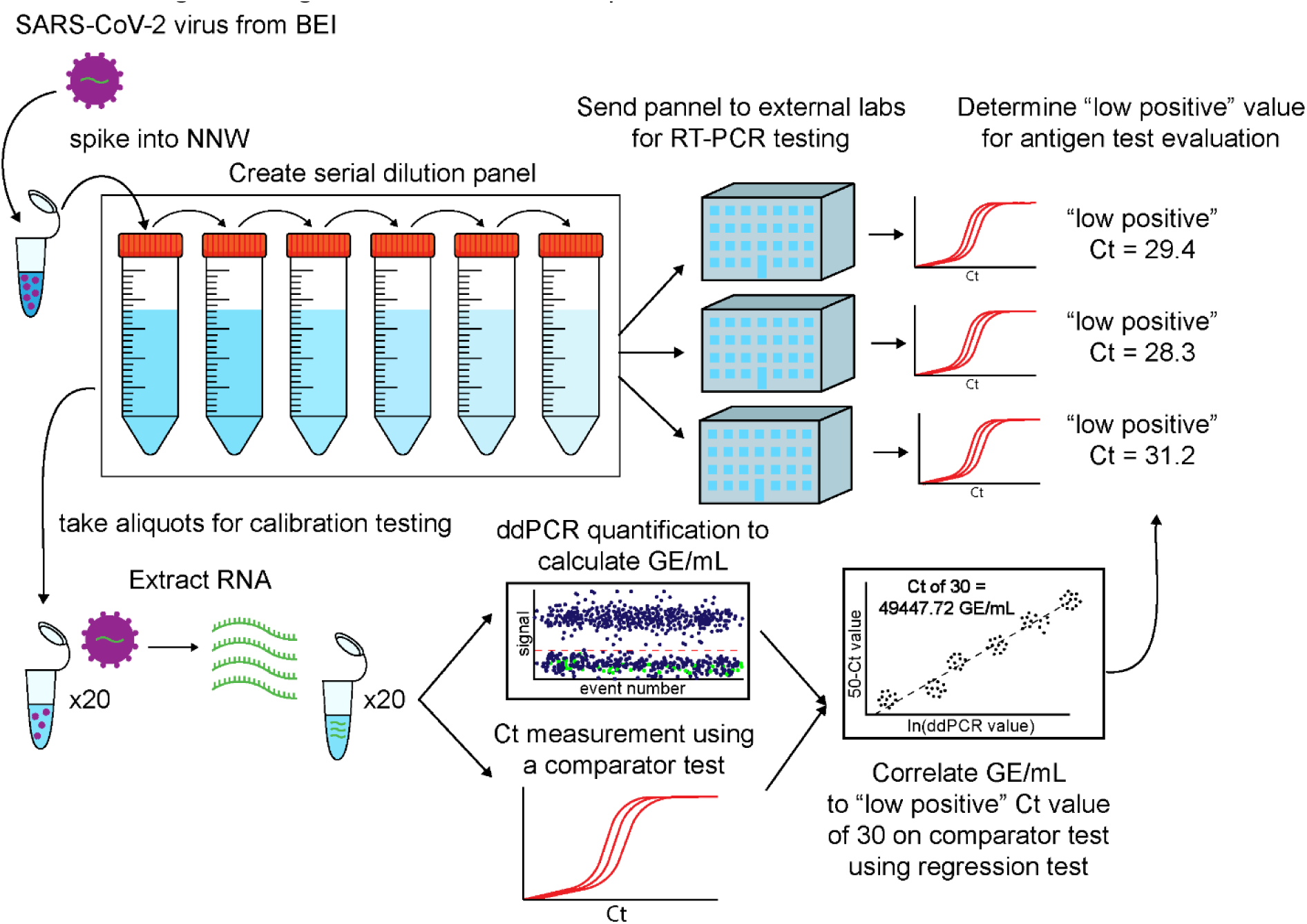
Workflow for generating a SARS-CoV-2 dilution panel and calibration curves for GE/mL to Ct value

To accomplish establishment of a standard, we first determined the GE/ml that corresponds to C_T_ =30 for the E Gene using the cobas® SARS-CoV-2-FluA/B at Tricore Reference Laboratories (Albuquerque, New Mexico). To do this, we used droplet digital PCR (ddPCR) a calibration-free method for absolute DNA quantitation.^29–31^ A C_T_ value of 30 was designated as the “low positive” cutoff as it was used in December 2021 for granting EUA for two Rapid Antigen Tests (RATs)-Osang and Xiamen Boson^23,24^. We determined by ddPCR, that the “low positive” cutoff (i.e., C_T_ 30) for the E gene using cobas® SARS-CoV-2-FluA/B at Tricore was 49447.72 GE/ml. We then determined the C_T_ value corresponding to this GE/ml standardized cutoff in two additional laboratories (cobas® SARS-CoV-2 test at Emory Medical Laboratory (EML) Services (Emory Healthcare, Atlanta, Georgia)), cobas® SARS-CoV-2 Test at Quest (Quest Diagnostics) and 2 additional assays (BD Max at Tricore Reference Laboratories and Cepheid Xpert Xpress CoV-2 plus at Emory/Children’s Laboratory for Innovative Assay Development (ELIAD)).

The method, procedure and statistical analysis described here and summarized in Figure 1 below, can now be used to provide a tool for standardization of all FDA acceptable comparators. We have used this protocol to calibrate the Roche cobas® 6800, BDMax and Cepheid in multiple clinical testing laboratories. We also calibrated the Center for Disease Control (CDC) assay, where we used the primer/probe sequence that had undergone functional testing for detection of SARS-CoV-2 at the CDC.

### Purpose of the Study

The purposes of this study are to 1) Define a “GE/mL standard” that corresponds to “low positive” cutoff (C_T_ =30 on the cobas® SARS-CoV-2-FluA/B assay at Tricore Reference Laboratory, 2) Determine the C_T_ value that corresponds to this “GE/mL standard cutoff” for each comparator assay, and 3) Generate calibration curves for GE/mL to C_T_ across multiple comparator assays, all done using the same heat inactivated BEI Wuhan strain and identical production lot.

### Study Design

A calibration curve was established by testing a series of two-fold dilutions of SARS-Related Coronavirus 2, (Isolate USA-WA1/2020, Heat Inactivated from Biodefense and Emerging Infections Research Resources Biorepository (BEI Resources), Catalog no. NR-52286, Lot number-70048020) spiked into SARS-CoV-2 negative pooled nasal cavity wash (NNW) (from Lee Biosolutions, Catalog number-991-26-P). Protocol details for preparing dilutions are described in Supplementary methods. Briefly, we used a 73-fold dilution of virus stock to generate Dilution 1. Further dilutions (total of 12) were generated by a series of 1:2 dilutions in NNW. RNA was isolated from twenty 140μl aliquots of each dilution. RNA was then used to determine N2 and SC2 C_T_ using CDC primers (primers with sequence identical to that established by CDC for RT-PCR testing) and was also used for ddPCR with N2 and SC2 primers/probe sets to generate GE/ml values (**Table 1**).

**Table 1:**
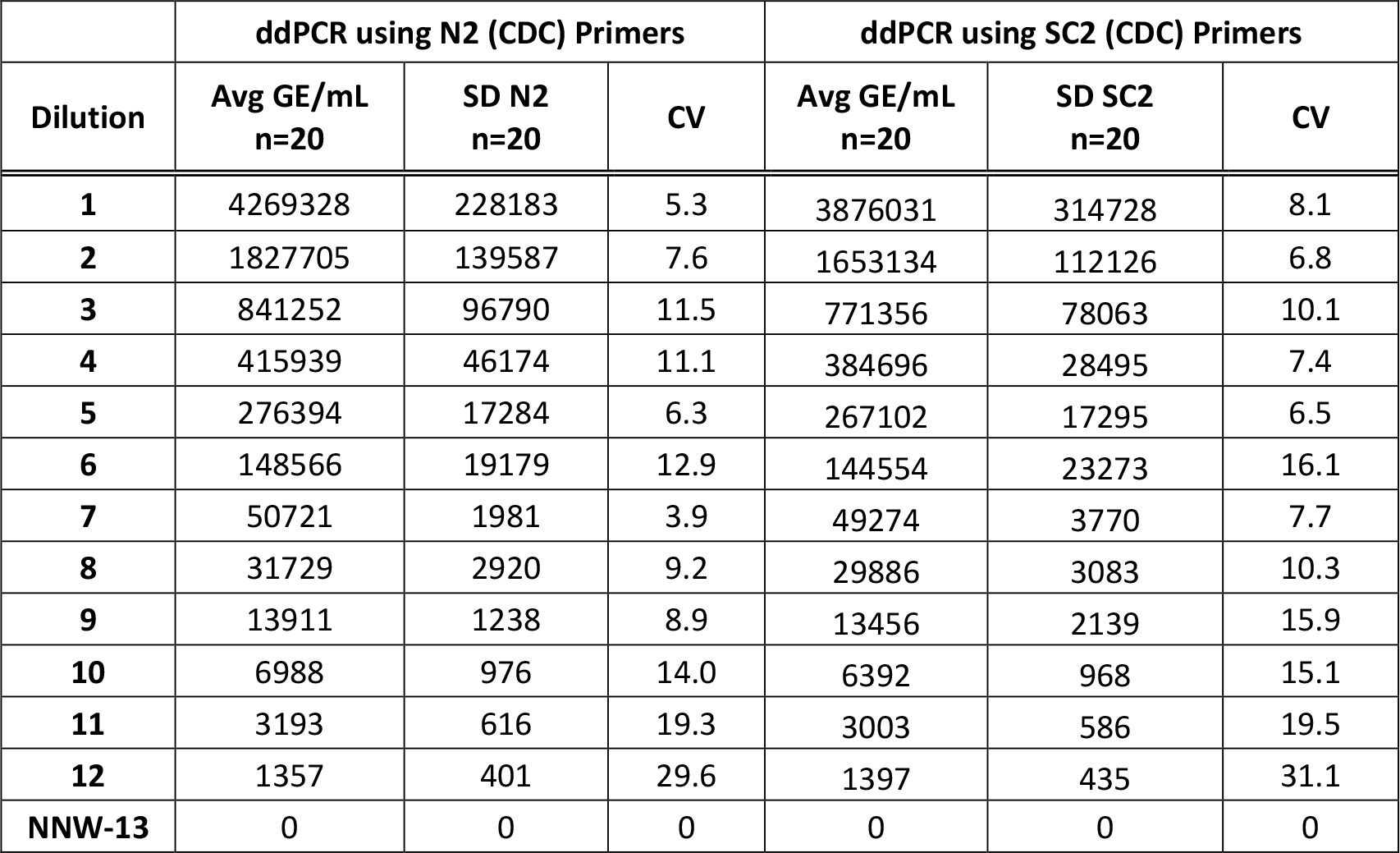
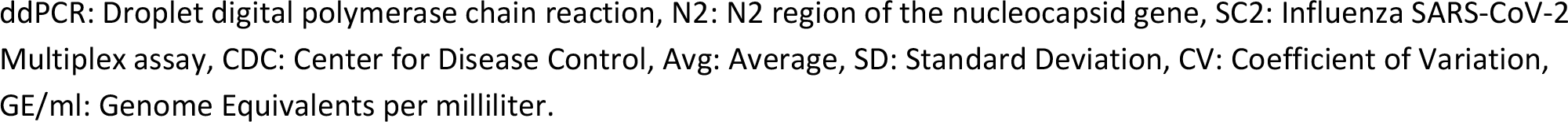
ddPCR values for 20 replicates using N2 and SC2 CDC primers. (Primary data with individual values is available in **supplementary data table S1 and S2**).

|  | ddPCR using N2 (CDC) Primers |  |  | ddPCR using SC2 (CDC) Primers |  |  |
| --- | --- | --- | --- | --- | --- | --- |
| Dilution | Avg GE/mL<br>n=20 | SD N2<br>n=20 | CV | Avg GE/mL<br>n=20 | SD SC2<br>n=20 | CV |
| 1 | 4269328 | 228183 | 5.3 | 3876031 | 314728 | 8.1 |
| 2 | 1827705 | 139587 | 7.6 | 1653134 | 112126 | 6.8 |
| 3 | 841252 | 96790 | 11.5 | 771356 | 78063 | 10.1 |
| 4 | 415939 | 46174 | 11.1 | 384696 | 28495 | 7.4 |
| 5 | 276394 | 17284 | 6.3 | 267102 | 17295 | 6.5 |
| 6 | 148566 | 19179 | 12.9 | 144554 | 23273 | 16.1 |
| 7 | 50721 | 1981 | 3.9 | 49274 | 3770 | 7.7 |
| 8 | 31729 | 2920 | 9.2 | 29886 | 3083 | 10.3 |
| 9 | 13911 | 1238 | 8.9 | 13456 | 2139 | 15.9 |
| 10 | 6988 | 976 | 14.0 | 6392 | 968 | 15.1 |
| 11 | 3193 | 616 | 19.3 | 3003 | 586 | 19.5 |
| 12 | 1357 | 401 | 29.6 | 1397 | 435 | 31.1 |
| NNW-13 | 0 | 0 | 0 | 0 | 0 | 0 |

ddPCR: Droplet digital polymerase chain reaction, N2: N2 region of the nucleocapsid gene, SC2: Influenza SARS-CoV-2 Multiplex assay, CDC: Center for Disease Control, Avg: Average, SD: Standard Deviation, CV: Coefficient of Variation, GE/ml: Genome Equivalents per milliliter.

Subsequently, the dilutions were tested in 20 replicates with FDA acceptable EUA comparator assays (cobas® 6800) in three different laboratories (EML, Tricore Reference laboratory and Quest Diagnostics), BD Max (Tricore) and Cepheid (ELIAD). Experimental controls were tested each time a sample set was analyzed. Controls (positive and negative) are run along with samples. The results from controls have to meet a preset criteria for passing, wherein the positive control should yield positive result and the negative control a negative result. This ensures that results from all samples included in that run are reliable. For a description of the Candidate Device, controls, including proposed intended use and indications for use, refer to the Quick reference guide (QRI).

Our goal was to create a dilution panel using heat inactivated SARS-CoV-2 Wuhan strain from BEI with C_T_ of the comparator ranging from ∼22 to ∼35 (12 dilutions, with dilution 13 being NNW) and ddPCR (N2, SC2 gene copy number denoted as GE/mL). The data were analyzed by the Passing Bablok and Deming regression models and were reported as the GE/ml at C_T_ = 30 of the comparator assays.

Samples sent to testing laboratories and CRO’s are blinded and include three unblinded controls in addition to the 12 blinded tubes containing virus dilutions. All data is submitted to Emory in excel format and then decoded by lead authors of this study who have the key for unblinding samples.

### Acceptance Criteria

Invalid results due to internal control failure require retesting. If the results are still invalid after re-testing three times, an invalid result is to be reported. Positive and Negative study controls should produce the expected results for the study to be valid.

## Materials and Methods

Detailed description of materials, methods and equipment needed are described in the supplement. Briefly, we first ensured that the nasal matrix used for virus dilution is indeed negative for the presence of SARS-CoV-2. RNA was isolated from an aliquot of NNW and used in a one step RT-PCR reaction to determine the presence of SARS-CoV-2 N2 gene. If the Ct for N2 is 0, then then that aliquot of nasal wash is deemed to be negative. After this, we used NNW to create a series of virus dilutions using heat inactivated SARS-CoV-2 from BEI resources. We then aliquoted appropriate amounts, as required for each individual comparator assay into 50ml falcon tubes and stored the samples at -80°C. We used twenty 140μl aliquots to isolate RNA, and to carry out the N2 and SC2 RTqPCR and ddPCR assays. Subsequently, aliquots of all 13 dilutions were distributed to various labs for analysis using cobas® 6800, BD Max and Cepheid. All samples that needed to be shipped were sent using overnight shipping on dry ice to ensure that they arrived frozen. Labs were instructed to thaw only the number of samples that could be tested in one day. After completion of testing, C_T_ values were provided to us (MG, AR, LB, RP and AW), and statistical analysis was performed by Emory University Pediatrics Statistics Core using methods approved by the FDA.

### Statistical Methods

The first aim of this study was to determine the GE/mL that corresponds to C_T_ =30 for the E Gene using the cobas®SARS-CoV-2-FluA/B assay data generated at Tricore Reference Laboratory. Once this was established, we determined the C_T_ value corresponding to C_T_ =30 (E Gene) across several comparator assays and multiple instruments.

We used Passing-Bablok and Deming regression to model the relationship between cobas® SARS-CoV-2-FluA/B, E gene calculated C_T_ value and ddPCR GE/ml because both the C_T_ and ddPCR values are measured with error which violates the assumptions needed to perform ordinary least squares linear regression. Passing-Bablok, which does not assume normality but does assume a strong linear correlation, was first fit to the data to identify outliers through visual inspection. After removing outliers identified from the Passing-Bablok procedure, Deming regression, which requires fewer assumptions about the relationship between the variables, was used to fit the data and determine ddPCR GE/ml for E Gene C_T_ value of 30 using the cobas® SARS-CoV-2-FluA/B at Tricore. To estimate the equivalent C_T_ from other instruments and laboratories, we performed a similar analysis. A Passing-Bablok model was first fit to find and remove outliers and then a Deming regression was applied to estimate the relationship between C_T_ value and ddPCR GE/ml. We then determined the C_T_ value for each instrument at the level of ddPCR GE/ml established above.

The delta parameter used in Deming regression was set equal to the average ratio of the variance between the natural-log transformed ddPCR and the 50 – C_T_ values (to transform the C_T_ into a positive slope) for each instrument. All analyses and graphs were conducted using the mcr package in R Version 4.2.0 (https://cran.r-project.org/web/packages/mcr/index.html).^32^

## Results

Dilutions of heat inactivated SARS-CoV-2 were prepared as described previously in NNW. Twenty 140μl aliquots of each dilution were used to isolate RNA, which was then used to determine GE/ml values of N2 gene using both N2 and SC2 primers by ddPCR as seen in **Table 1**. NNW (NNW-13) was also analyzed by ddPCR using both N2 and SC2 primers, to verify value of 0.

Next, samples were sent to TriCore Reference Laboratory for analysis using the cobas® 6800 where the SARS-CoV-2 & Influenza A/B Assay was used to generate C_T_ values for ORF1a, and E gene as seen in **Table 2**. Each dilution was analyzed in replicates of 20. One tube contained NNW and was used as a negative control (NNW-13) and was also analyzed in replicates of 20.

**Table 2:** ORF1a and E gene C_T_ values obtained for 20 replicates from Roche Cobas SARS-CoV-2 & Influenza A/B Assay by TriCore Reference Laboratories. R^2^ was calculated in excel with dilution number on y-axis and C_T_ on x-axis, in a scatterplot. (Primary data with individual C_T_ values used in Passing Bablok and Deming Regression analysis is available in **supplementary data table S3**)

|  | Roche Cobas ORF1a (TriCore) |  |  | Roche Cobas E (TriCore) |  |  |
| --- | --- | --- | --- | --- | --- | --- |
| Dilution | ORF1a-C<sub>T</sub> Avg | C<sub>T</sub> -SD | CV | E- C<sub>T</sub> - Avg | C<sub>T</sub> -SD | CV |
| 1 | 22.9 | 0.2 | 1.0 | 23.5 | 0.2 | 0.7 |
| 2 | 25.3 | 0.2 | 0.6 | 26.0 | 0.1 | 0.6 |
| 3 | 26.0 | 0.2 | 0.6 | 26.7 | 0.2 | 0.7 |
| 4 | 27.1 | 0.1 | 0.4 | 27.7 | 0.1 | 0.4 |
| 5 | 28.4 | 0.3 | 0.9 | 28.8 | 0.3 | 0.9 |
| 6 | 27.9 | 0.2 | 0.5 | 28.2 | 0.6 | 2.2 |
| 7 | 30.8 | 0.2 | 0.5 | 30.7 | 0.2 | 0.6 |
| 8 | 30.1 | 0.2 | 0.5 | 30.1 | 0.1 | 0.5 |
| 9 | 32.2 | 0.2 | 0.7 | 32.0 | 0.2 | 0.6 |
| 10 | 31.9 | 0.2 | 0.5 | 31.7 | 0.1 | 0.5 |
| 11 | 33.0 | 0.1 | 0.4 | 32.7 | 0.1 | 0.4 |
| 12 | 34.2 | 0.2 | 0.7 | 33.8 | 0.2 | 0.6 |
| R<sup>2</sup> | 0.9307 |  |  | 0.9052 |  |  |
| NNW-13 | 0.0 |  |  | 0.0 |  |  |

ORF1a: Open Reading Frame 1a, E: Envelope, C<sub>T</sub>: Cycle threshold, NNW: Negative Nasal Wash, Avg: Average, SD: Standard Deviation, CV: Coefficient of Variation, R<sup>2</sup>: r squared

**Table 3.** Calculations of variance and standard deviation ratios between the variance of the log-transformed ddPCR GE/ml values and the transformed Roche E2 C_T_ values along with their respective averages.

| Dilution | Var<sup>a</sup> X:<br>Var(Log(ddPCR)) | Var Y:<br>Var(50 - (Roche cobas® E C<sub>T</sub>)) | Delta:<br>Var(Y)/Var(X) | CV<sup>b</sup>(Y)/CV(X) |
| --- | --- | --- | --- | --- |
| 2 | 0.01 | 0.02 | 3.48 | 1.87 |
| 3 | 0.01 | 0.03 | 2.38 | 1.54 |
| 4 | 0.01 | 0.01 | 1.04 | 1.02 |
| 5 | 0 | 0.07 | 17.73 | 4.21 |
| 6 | 0.02 | 0.02 | 1.44 | 1.20 |
| 7 | 0 | 0.04 | 24.04 | 4.90 |
| 8 | 0.01 | 0.02 | 2.31 | 1.52 |
| 9 | 0.01 | 0.03 | 4.03 | 2.01 |
| 10 | 0.02 | 0.02 | 1.09 | 1.05 |
| 11 | 0.04 | 0.02 | 0.43 | 0.66 |
| 12 | 0.1 | 0.05 | 0.44 | 0.67 |
| Average | 0.02 | 0.03 | 5.31 | 1.88 |

<sup>a</sup>Var: Variance, <sup>b</sup>CV: coefficient of variance

Using the above values, the ddPCR value corresponding to cobas® 6800 E C_T_ of 30 was calculated using Deming Regression as described below. This procedure was repeated for each instrument, assay, and gene using the ddPCR values obtained from each sample in the dilution series. Every time a new dilution series is prepared, the ddPCR values obtained for that series will help determine low-positive cutoff.

When performing Passing-Bablok Regression using 50 – cobas® 6800 E C_T_ on the y-axis and natural log transformed ddPCR GE/ml on the x-axis, we see in Figure 2 that one sample and one dilution, circled in red in the plot were considered outliers and were to be removed before performing Deming regression. Outliers were identified and removed by visual inspection, and the plot after outliers were removed is shown in **Figure S1**. As the Passing-Bablok regression method is robust to outliers^33^, there is no concern that the outlier cluster circled in red on the top-right of **Figure 2** will violate linearity.

**Figure 2.**
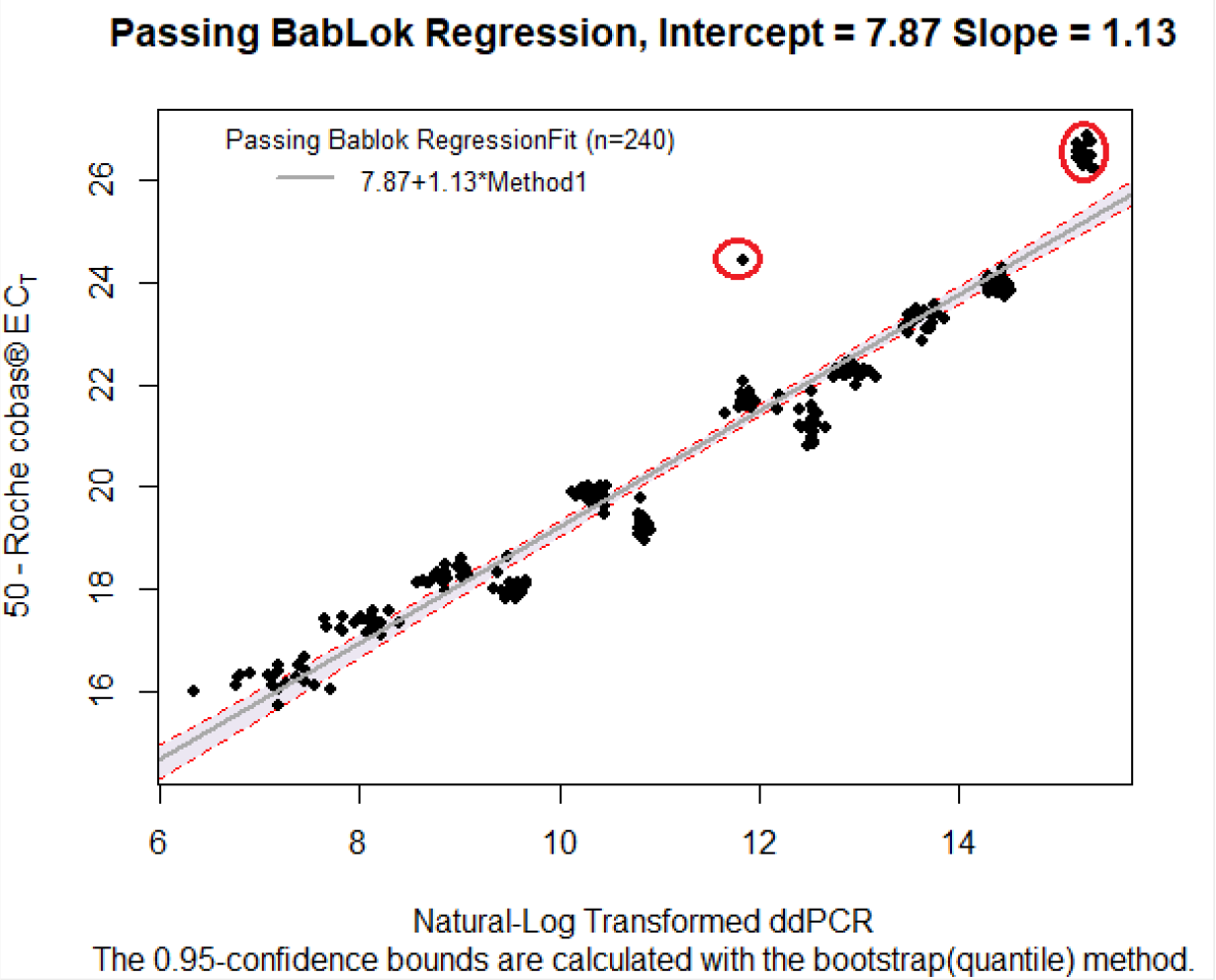
Passing-Bablok Regression with log-transformed ddPCR GE/ml concentration on the x-axis and 50 – Roche E C_T_ on the y-axis. Outliers to be removed are denoted with red circles.

**Figure 3.**
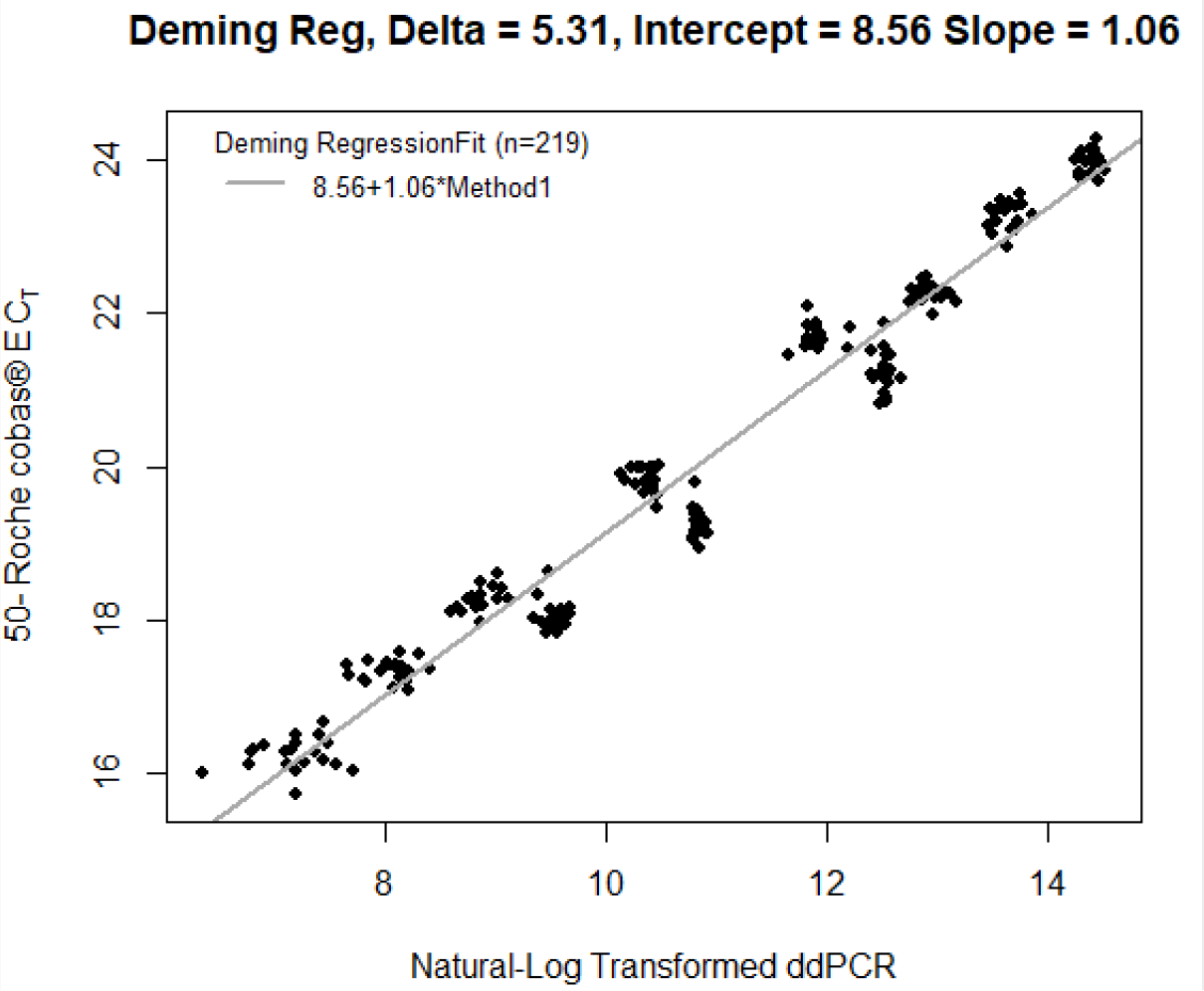
Deming Regression of Data without outliers with log-transformed ddPCR GE/ml on the x-axis and 50 – Roche cobas® E C_T_ on the y-axis, using the computed average ratio of the variances as our delta.

The table below tells us that the delta parameter (calculated as the average ratio of the variance of the Roche cobas® E C_T_ to the variance of the natural-log transformed ddPCR values across every dilution) we should use for the Deming regression is 5.31. Dilutions 5 and 7 had variance ratios that were considerably high compared to the others, with values of 17.73 and 24.04 respectively.

From the Deming regression that was performed and plotted below, we would expect that a 1-unit increase in natural log ddPCR GE/ml value leads to approximately a 1.06 unit increase in cobas® E C_T_ value.

Using these parameters from the Deming regression method, we found that the ddPCR value that corresponds to a 30 C_T_ on the cobas® instrument at Tricore with the E gene target is **49447**.**72 GE/mL**.

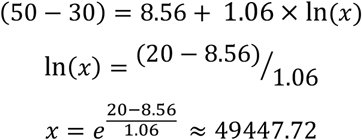

This value of 49,447.72 is the baseline ddPCR that we will use to calculate the 30 C_T_ equivalent for all other instruments, assays, and genes. For example, for the TriCore BD Max N2 instrument, we calculate its 30 CT equivalent as follows:

1. Generate Passing-Bablok regression model (**Figure S3**)
2. Visually inspect and remove outliers (**Figure S5**)
3. Generate variance ratios as required by Deming regression model (**Table S14**)
4. Generate Deming regression model (**Figure S7**)
5. Using parameters produced by the Deming regression and the baseline of 49,447.72 GE/ml, generate the equivalent C_T_

We followed this same procedure for three more instruments: TriCore BD Max N1, Emory CDC N2, and Emory CDC SC2. The supplement shows:

1. The generated Passing-Bablok regression model plots (see **Figure S2** for TriCore BD Max N1, **Figure S8** for Emory CDC N2, and **Figure S9** for Emory CDC SC2).
2. The Passing-Bablok model plots with any outliers removed (see **Figure S4** for TriCore BD Max N1 and **Figure S10** for Emory CDC SC2).
3. The compiled variance ratios needed for Deming regression (see **Table S14** for TriCore BD Max N1/Tricore BD Max N2 and **Table S15** for Emory CDC N2/Emory CDC SC2).
4. The generated Deming regression model plots (see **Figure S6** for TriCore BD Max N1, **Figure S11** for Emory CDC N2 and **Figure S12** for Emory CDC SC2).
5. The calibrated C_T_ values for each instrument using the baseline of 49,447.72 GE/ml.

### Tricore BD Max N2 Example Calculation

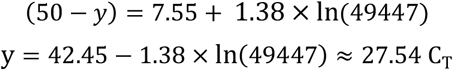

For each new production lot of an assay, we can use the above formula to calibrate Ct values back to the original lot given the Ct and ddPCR values produced by the new lot. We do not expect much variance in calibrated Ct value calculations as production lot changes, nor do we expect a need to change the formula with different variants of SARS-CoV-2 so long as new mutations do not greatly affect primer binding.

As described earlier, dilutions were tested in replicates of twenty with FDA EUA acceptable comparator assays (cobas®6800) in three different laboratories (Emory Medical Laboratory, TriCore Reference laboratory and Quest Diagnostics), BD Max (TriCore) and Cepheid (ELIAD). Average and individual C_T_ for all genes, and for all instruments are available in supplementary tables (**Tables S4** to **S13**).

#### Comparing Calibrated C_T_ Value Calculations Across Different Instruments

From observing the table below, we see that all calibrated C_T_ values are between 26-31, with Tricore cobas® Orf1a having the closest resemblance to the cobas® E 30 C_T_ target with a calibrated C_T_ value of 29.77 (**Table 2, Table S3**). The cobas® at both Emory Medical Laboratory (**Table S4** and **S5**) and Quest Diagnostics (**Table S6** and **S7**) performed similar to the cobas® platform at TriCore, with calibrated C_T_ values between 28.27 and 30.53. Calibrated C_T_ for BD Max N1 and N2 were within 1 C_T_ (**Table S8** and **S9**). Calibrated values for Cepheid Xpert Xpress CoV-2 plus showed the most variation and ranged from 27.79 for the E gene and 31.52 for the N2 gene (**Table S10** and **S11**). The N2 and SC2 C_T_ determined from RNA isolation and using CDC’s primers were very close to each other, denoting that both primer sets are equivalent in this context (**Table S12** and **S13**).

## Discussion

As of June 2023, there are approximately 300 molecular (nucleic acid-based) and over 50 rapid antigen tests that have received FDA EUA authorizations. Furthermore, a subset of the molecular assays, primarily PCR, have been designated as FDA EUA acceptable comparators for validating new diagnostic tests. However, it has been difficult to compare the performance of these tests by Positive or Negative Predictive Agreement (PPA & NPA) due to the differences in comparators used in various clinical trials and testing laboratories. In addition, it has been demonstrated that different testing laboratories will generate different C_T_ values using the same platform and identical samples due to slight differences in laboratory specific protocols and processing.^33^ The requirement of inclusion of at least 10% “low positive” samples in the clinical trials for rapid antigen test further complicates the correlation between molecular and antigen tests and among different rapid antigen tests.

When SARS-CoV-2 RNA levels have a low C_T_ value, this indicates that the clinical sample likely contains infectious viruses that are replicating and expressing proteins.^33,34^ However, high C_T_ values (>30) do not always indicate the presence of subgenomic RNA necessary for viral replication and structural protein (antigen) expression.^33^ It’s important to note that low C_T_ values often correlate positively with positive RATs. However, when the C_T_ value exceeds 30, it’s common for antigens to be absent in the clinical sample. This doesn’t mean that RATs are insensitive, but that the structure antigen is lacking in the sample.

While the same comparator (e.g. cobas®assay) might yield a different C_T_ with the same GE/ml (at 49,447), the protocol described in this report provides a method to standardize all tests and testing laboratories to a single standard (i.e. Genome Copies per mL; GE/mL). This produces a standard of PCR calibration regarding COVID-19 samples and allows us to directly compare the C_T_ values of different testing platforms and laboratories, greatly advancing our ability to compare all the EUA authorized tests and to establish an uniform performance acceptance criteria for future EUA authorizations. Working closely with the FDA, we have used this protocol to establish the low positive cutoff using the Roche 6800 COVID assay E gene to be 49,447 GE/mL at the TriCore Laboratories. This GE/mL cutoff was used as the low positive cutoff for two of the EUAs (Osang and Xiamen Boson).^23,24^ We have since calibrated a number of FDA EUA acceptable comparator platforms and in different clinical testing laboratories (**Table 4**). Additionally, this method of low positive cuff off was used for granting EUA to other RATs, namely: Genbody, watmind, Azure Pen, Mologic and Ellume.

**Table 4.**
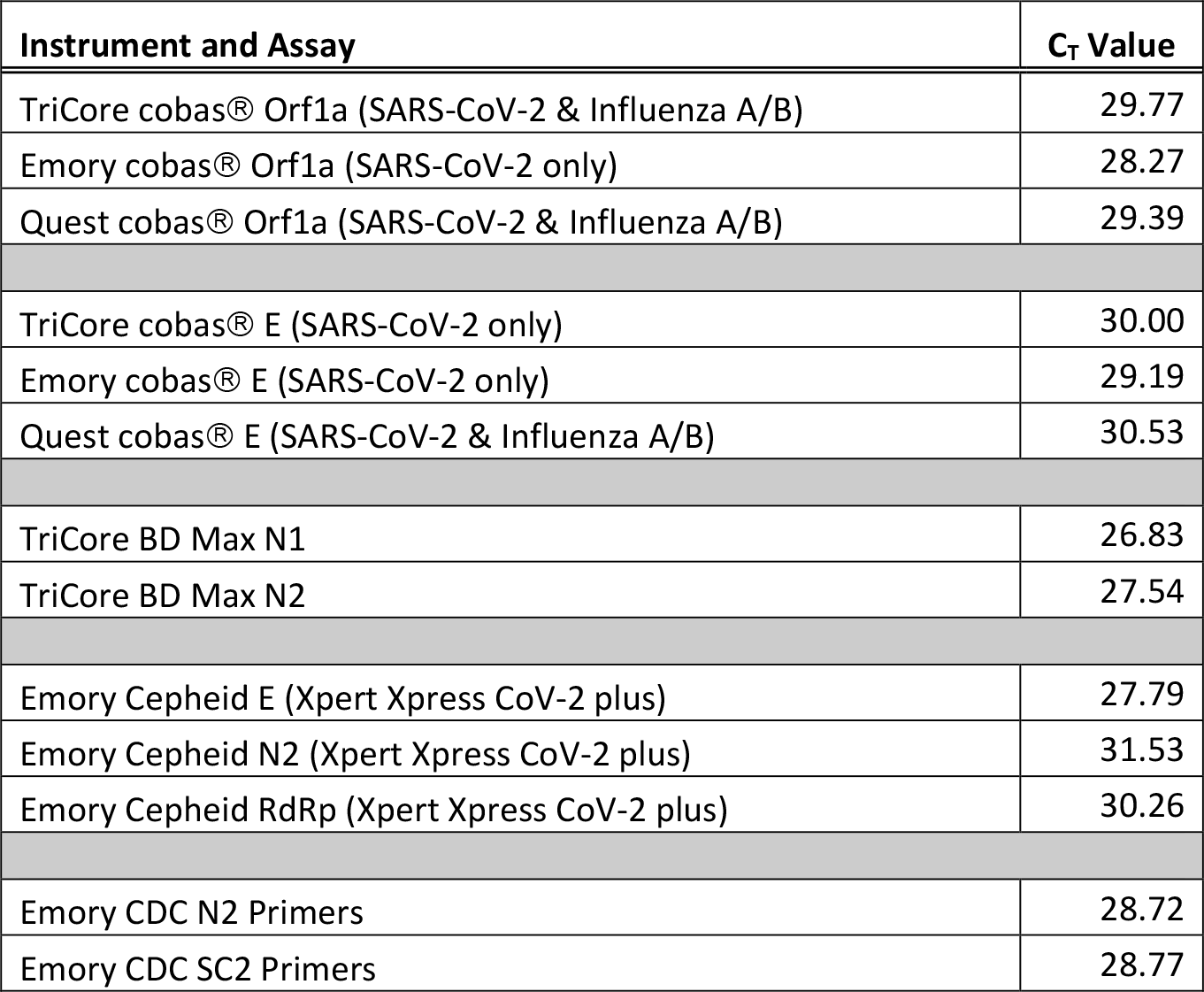
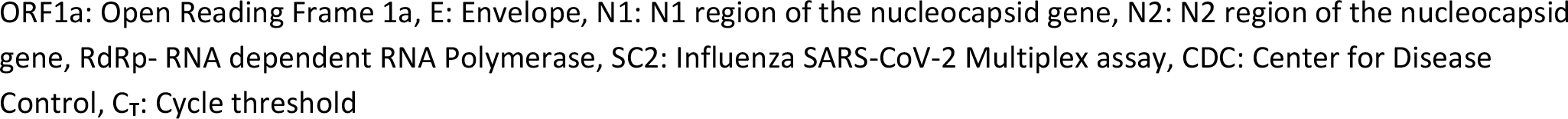
Computed calibrated C_T_ cutoff value for low positive samples for each of 9 instruments at 49,447 GE/mL (equivalent to cobas® at Tricore Laboratory used for EUA authorizations).

| Instrument and Assay | $C_T$ Value |
| --- | --- |
| TriCore cobas® Orf1a (SARS-CoV-2 & Influenza A/B) | 29.77 |
| Emory cobas® Orf1a (SARS-CoV-2 only) | 28.27 |
| Quest cobas® Orf1a (SARS-CoV-2 & Influenza A/B) | 29.39 |
| TriCore cobas® E (SARS-CoV-2 only) | 30.00 |
| Emory cobas® E (SARS-CoV-2 only) | 29.19 |
| Quest cobas® E (SARS-CoV-2 & Influenza A/B) | 30.53 |
| TriCore BD Max N1 | 26.83 |
| TriCore BD Max N2 | 27.54 |
| Emory Cepheid E (Xpert Xpress CoV-2 plus) | 27.79 |
| Emory Cepheid N2 (Xpert Xpress CoV-2 plus) | 31.53 |
| Emory Cepheid RdRp (Xpert Xpress CoV-2 plus) | 30.26 |
| Emory CDC N2 Primers | 28.72 |
| Emory CDC SC2 Primers | 28.77 |

ORF1a: Open Reading Frame 1a, E: Envelope, N1: N1 region of the nucleocapsid gene, N2: N2 region of the nucleocapsid gene, RdRp- RNA dependent RNA Polymerase, SC2: Influenza SARS-CoV-2 Multiplex assay, CDC: Center for Disease Control, C<sub>T</sub>: Cycle threshold

As and when assays need to be calibrated, new dilutions in NNW with the the same heat inactivated virus as used in the current study can be prepared according to the methods described here. The RT-PCR and GE/mL of each dilution will be determined using an N2 primer/probe mix in replicates of 20 for internal verification that dilutions are accurate and that RT-PCR Ct increase and ddPCR GE/ml values decrease as with dilution. Dilutions will be sent to testing laboratories on dry ice, and C_T_ values will be determined. Then, using the 49,447.72 GE/mL baseline we calculated earlier, the C_T_ 30 equivalent for these new assays will be calculated. This resulting C_T_ value can then be used as a threshold for low positive cut-off for these assays.

### Limitations of the Study

Although this study establishes a protocol for standardization using GE/mL and an absolute cut-off value for “low positive” samples for evaluation of antigen tests, this method does not account for the variation of clinical samples. Studies have shown that C_T_ value and GE/mL can not be used to determine infectiousness.^35^ Futhermore, C_T_ values have a different kinetics over the course of infection compared to antigen levels, and the kinetics may change based on hard to predict external factors such as immunization status, viral variant characterstics, and treatments received. Thus, regulatory approval should therefore also include validation using clinical patient samples, as well as using the standardized samples described here.

Finally, the cut-off Ct value of 30 is an arbitrary standard, and may be adjusted in light of additional data or changes in virus characteristics. In that case, it will be possible to recalculate the GE/ml and recalibrate with the comparators using the methods described here.

## Conclusion

Here we have developed a novel methodology and applied it to calibrate and compare different FDA EUA acceptable comparator instruments from different manufacturers and laboratories. This method has been used in seven EUA applications granted as of September 28^th^ 2023. The procedure described here therefore serves as an instruction manual for calibrating the user’s desired clinical comparator assay in the precise manner acceptable to the US FDA and interpreting the results of the calibration. As such, this paper provides the conceptual framework and procedures that can be followed for any new device seeking EUA status. All clinical data submitted to the FDA for new SARS-CoV-2 diagnostics will be impacted by this important work.

## Data Availability

Data and code are available at: https://github.com/rbparso/Calibration-Curve

## Data and Code Availability

Data and code are available at: https://github.com/rbparso/Calibration-Curve

## Acknowledgments

We thank Biodefense and Emerging Infections Research Resources Biorepository (BEI Resources) for providing us with SARS-Related Coronavirus 2, Isolate USA-WA1/2020, Heat Inactivated, Catalog no. NR-52286, Lot number-70048020 that was used in this study. We thank the Food and Drug Administration and Dr. Yi Yan for advice regarding the statistical methods used in the paper. We thank Emory Molecular Lab for analyzing samples with cobas® 6800, TriCore Reference Laboratories for analyzing samples with cobas® 6800 and BD MAX, and Quest Diagnostics along with University of Massachusetts and Dr. Apruv Soni analyzing samples with cobas® 6800.

## Author contributions

Conceptualization: EL, AR, LB, MG

Methodology: AR, LB, MG, EL, RP, AW, HBB, WOS, JAS, KM, TB, MS

Investigation: AR, LB, MG, EL, RP, AW, HBB, WOS, JAS, KM, TB, MS

Funding: WAL

Project administration: AR, LB, MG, EL

Supervision: AR, LB, MG, EL

Writing – original draft: MG, EL, RP, AR, LB, AW

Writing – review & editing: AR, JL, RP, MG, AW, LB, EL, WAL

**Declaration of interests: Authors declare that they have no competing interests**.

