## Supplement for "Standardization and Comparison of Emergency Use Authorized COVID-19 Assays and Testing Laboratories"

**Supplementary Methods:**

*NOTES: This protocol describes the expected study procedure. Modifications are allowed to accommodate specific study objectives or circumstances provided they are approved by the Principal Investigator. All changes and rationale should be documented on the appropriate forms.*

*All work should be performed in a dedicated Biosafety Cabinet* (*BSC).*

*Run external controls once per new user and with new cartridge/cassette lots. Samples can be run in batches of 5-10 for convenience as allowed by the procedure.*

**Equipment and Materials to be provided for Device being analyzed:**

- Complete Candidate Device test kit per the QRI.
- External positive and negative controls to be provided by the device being tested.

**Equipment and Materials Provided by labs at Emory, Contract Research Organization or other:**

- Calibrated Pipettes (Gilson, Rainin LTS P-2, P-20, P-200, P-1000 or equivalent)
- Vortex
- Biosafety Cabinet
- Refrigerator, 2 - 8ºC
- Freezer, ≤-70°C
- Timer
- Bleach (10% for cleaning, 50% for liquid decontamination)
- 70% Isopropanol
- Personal Protective Equipment
- Aerosol barrier filter, nuclease-free pipette tips
- Microfuge tubes (for preparing dilutions)
- Negative Nasal Cavity Wash from Pooled Human Donors (NNW) (Lee Biosolutions, 991-26-P)

**Reagents, Consumables, and Equipment Used for Calibration Curve**

- Calibrated Pipettes (Gilson, Rainin LTS P-2, P-20, P-200, P-1000 or equivalent)
- Nasal Cavity Wash from Pooled Human Donors (Lee BioSolutions – Catalog number: 991-26-P
- SARS-Related Coronavirus 2, Isolate USA-WA1/2020, Heat Inactivated (BEI, Catalog no. NR- 52286, Lot number 70048020)
- MagMAX Viral RNA Isolation Kit (Applied Biosystems by Thermo Fisher Scientific - Catalog number: AM1939)
- KingFisher Apex system (Thermo Fisher Scientific - Catalog number: 5400920)
- qScript XLT 1-Step RT-qPCR ToughMix (QuantaBio/VWR - Catalog numbers: 95132/76047-080)
- 2019-nCoV Kit, 500 reaction combined (N2 primers/probe set) [Integrated DNA Technologies (IDT)], Catalog numbers: 592223/10006606)
- Quantitative Synthetic RNA from SARS (positive control)—Related Coronavirus 2 (BEI – Catalog/lot numbers NR-52358/70035241)
- RNase-free water (Qiagen – Catalog number – 166034309)
- SC2 Forward Primer (IDT - Catalog number: 10011565)
- SC2 Reverse Primer (IDT - Catalog number: 10011566)
- SC2 (TexRd) Probe (IDT - Catalog number: 10011567/70035241)
- SC2 (HEX) Probe (IDT – Catalog number/lot number: 329293312/62790567)
- One-Step RT-ddPCR Advanced Kit for Probes (BioRAD – Catalog number: 1864022)
- DTT (BioRAD – Catalog number: 64460539)
- Supermix (BioRAD – Catalog number: 64464141)
- Reverse transcriptase (BioRAD – Catalog number: 64464920)
- ddPCR Buffer Control Kit for Probes: Negative control (BioRAD – Catalog number: 1863052)
- Positive control (SC2) - CDC Influenza SARS-CoV-2 Multiplex assay positive controls kit, FluSC2PC-EUA (ATCC – catalog number: 200811A/Part# MR 669)
- LightCycler 480 Instrument II (Roche – Catalog number: 04911016001)
- QX200 Auto DG Droplet Digital PCR System Auto DG Droplet Digital PCR System for EvaGreen or probe-based digital PCR applications, includes Automated Droplet Generator, QX200 Droplet Reader, computer, and QuantaSoft Analysis Pro Software (BioRAD – Catalog number: 1864100)
- PX1 PCR Plate Sealer PCR plate sealer includes heat-sealing instrument, plate support block that holds 96-well and 384-well plates, sealing frame, power cord (BioRAD – Catalog number: 1814000)
- Automated Droplet Generator Oil for Probes (BioRAD - Catalog number: 1864110)
- C1000 Touch™ Thermal Cycler with 96–Deep Well Reaction Module Modular thermal cycler platform includes C1000 Touch Thermal Cycler chassis, 96--deep well reaction module, USB flash drive (BioRAD – Catalog number: 1851197)
- ddPCR™ Droplet Reader Oil2 L (2 x 1 L), oil for use with droplet reader in the QX200™/QX100™ Droplet Digital™ PCR Systems (BioRAD – Catalog number: 1863004)
- DG32 Automated Droplet Generator Cartridges (BioRAD – Catalog number: 1864108)
- ddPCR 96-Well Plates (BioRAD – Catalog number: 12001925)
- Pipet Tips for the AutoDG System (BioRAD – Catalog number: 1864120)
- Pipet Tip Waste Bins for the AutoDG System (BioRAD – Catalog number: 1864125)
- PCR Plate Heat Seal, Foil, Pierceable (BioRAD – Catalog number: 1814040)

1. Quality control of matrix: Nasal Cavity Wash from Pooled Human Donors (Lee BioSolutions – Catalog number: 991-26-P, Lot number 02H6571) is used as the matrix to prepare dilutions. It is first confirmed to be negative for SARS-CoV-2 using cobas®SARS-CoV-2 Test at Emory Molecular Labs and RNA isolation and N2 C_T_ determination as described here. The product arrives on dry ice in 50ml aliquots. A 50ml bottle is completely thawed. Contents are transferred to a 50ml falcon tube and briefly centrifuged at ~1000 rpm for 5-10 minutes to remove debris. Then, one 140μL aliquot is removed from each falcon tube, RNA is isolated and assayed in triplicate for the presence of the N2 gene of SARS-CoV-2 as described next. After confirming the lack of N2 C_T_ (C_T_=0) and negative by Roche Cobas® 6800, the required number of bottles are thawed, pooled, and used to make dilutions. SARS-CoV-2 negative Nasal Cavity Wash is denoted as Pooled Negative Nasal Cavity Wash (NNW).
2. Dilutions and analysis with 20 replicates: A dilution series of heat inactivated wild type SARS-Related Coronavirus 2 differing by 1 C_T_ from N2 C_T_ of ~22 till ~35 was prepared. To achieve this, SARS CoV-2 isolate USA-WA1/2020, Heat Inactivated (BEI, Catalog no. NR-52286, Lot number- 70033548) was used. The stock concentration of this lot of virus is 1.6X10^9^ GE/ml and 1.6X10^5^ TCID50/ml. This stock virus is diluted 73.3-fold in NNW to obtain the first dilution. Subsequently a series of 1:2 dilutions for 12 total dilutions in NNW were made. After completion of the dilution series, aliquots with volume sufficient for testing at 20 replicates for each type of test were made and stored at -80°C for long term storage (>1 week) or -20°C for short term storage (< 1 week).
3. Twenty 140µL aliquots of each dilution were used to isolate RNA (described below) and to carry out the N2 and SC2 RTqPCR and ddPCR assays (**Tables 1, S1 and S2**). For the CDC N2 and SC2 assays, negative control is NNW, while synthetic RNA SARS- CoV-2 control (BEI) is the positive control.
4. Samples were analyzed at three additional laboratories [cobas® SARS-CoV-2-FluA/B at TriCore (**Tables 2 and S3**)], cobas® SARS-CoV-2 Test at Emory Medical Laboratory (**Tables S4 and S5**), and cobas® SARS-CoV-2 Test by Quest for University of Massachusetts Laboratories (**Tables S6 and S7**] and 2 additional assays [BD Max at TriCore (**Tables S8 and S9**) and Cepheid Xpert Xpress CoV-2 plus at Emory/Children’s Laboratory for Innovative Assay Development (ELIAD) (**Tables S10 and S11**).
5. RNA Purification, RT-qPCR, and droplet digital PCR (ddPCR) Methods: Total RNA is extracted from twenty 140µL aliquots of each dilution, using MagMax Viral RNA Isolation Kit (Applied Biosystems) in a KingFisher Apex system (Thermo Fisher Scientific). RNA is eluted in 60µl, and one aliquot of 5µl of the eluate is used in a 10µl one-step RT-PCR reaction and reverse transcribed into cDNA with qScript XLT one-Step RT-PCR ToughMix (QuantaBio) using 2019-nCoV CDC EUA Kit, Primer/Probe Mix (N2 gene) CDC SC2 primers/probe mix in a LightCycler 480 II instrument (Roche) (**Tables S12 and S13**). Another aliquot of 5µL purified RNA is used in a 22µL ddPCR reaction with One-Step RT-ddPCR Advanced Kit for Probes BioRAD and IDT Primer/Probe Mix (N2 gene) and SC2 primer/probe mix (HEX SC2 Probe in a QX200 Droplet Digital PCR System (BioRad). Results of ddPCR are expressed in GE/ml (Tables 1, S1 and S2). Detailed standard operation protocols are cited on ***(2) Related Form(s) and Documents no. 2-7***).

*Reagents, Consumables, and Equipment Used for Calibration Curve Analysis performed at TriCore Reference Laboratories, Emory Medical Laboratory, and Quest Diagnostics for University of Massachusetts according to their protocols*

6. cobas®6800 (SARS-CoV-2 & Influenza A/B Assay): Testing of the panel (13 dilutions- differing by ~1 C_T_, and 20 replicates per dilution), was conducted using cobas® SARS-CoV-2 & Influenza A/B Assay by TriCore Reference Laboratories according to their protocol. (Average data in **Table 2**, with individual 20 replicate values in **Table S3**).
7. cobas® 6800 (Covid only): Testing of the panel (13 dilutions- differing by ~1 C_T_, and 20 replicates per dilution) at Emory Molecular Laboratory on the cobas® 6800 System was conducted according to the IFU for Roche cobas® 6800. (Average ORF1a, E and RNAse P gene values obtained from 20 replicates is shown in **Table S4**, with individual 20 replicate values in **Table S5**).
8. cobas® 6800 (SARS-CoV-2 & Influenza A/B Assay): Testing of the panel (13 dilutions- differing by ~1 C_T_, and 20 replicates per dilution), was conducted using cobas® SARS-CoV-2 & Influenza A/B Assay by Quest Diagnostics for according to their protocol. (Average ORF1a and E gene values obtained from is in Table S6, with individual 20 replicate values in **Table S7**).
9. BD MAX System: Testing of the panel (13 dilutions- differing by ~1 C_T_, and 20 replicates per dilution), was conducted using BD MAX System by TriCore Reference Laboratories according to their protocol. (Average N1, N2 and RNASeP gene values obtained from 20 replicates using BD MAX System is in **Table S8**, with individual 20 replicate values in **Table S9**).
10. Xpert Xpress CoV-2 plus by Cepheid: Testing of the panel (13 dilutions- differing by ~1 C_T_, and 20 replicates per dilution), was conducted using Xpert Xpress CoV-2 Plus Kit by Cepheid was be performed at ELIAD according to IFU (Average E, N2 and RdRp gene values for 20 replicates is in **Table S10**, with individual 20 replicate values in **Table S11**).

**SUPPLEMENTARY DATA**

**Table S1**: ddPCR values for each of the 20 replicates using the N2 CDC primer probe set (submitted as Table S1.csv file).

**Table S2**: ddPCR values for each of the 20 replicates using the SC2 CDC primer probe set (submitted as Table S2. csv file).

**Table S3**: ORF1a, and E gene values for each of the 20 replicates obtained from cobas® SARS-CoV-2-FluA/B at Tricore cobas®6800 (submitted as Table S3. csv file).

**Table S4**: Average of ORF1a, E and RNAse P gene values obtained from 20 replicates from cobas®6800 (Covid only by Emory Molecular Laboratories. R^2^ was calculated in excel with dilution number on y-axis and C_T_ on x-axis, in a scatterplot.

|  | **Roche Cobas ORF1a** | | | **Roche Cobas E** | | | | | **Roche Cobas RNaseP** | | |
| --- | --- | --- | --- | --- | --- | --- | --- | --- | --- | --- | --- |
| **Dilution** | **ORF1a C_T_ Avg** | **C_T_ SD** | **CV** | **E C_T_ Avg** | **C_T_ SD** | **CV** | | **RNaseP C_T_ Avg** | | **C_T_ SD** | **CV** |
| **1** | 22.1 | 0.22 | 0.99 | 22.9 | 0.30 | 1.29 | | 33.4 | | 0.24 | 0.71 |
| **2** | 23.2 | 0.15 | 0.63 | 24.0 | 0.17 | 0.70 | | 33.3 | | 0.22 | 0.67 |
| **3** | 24.3 | 0.18 | 0.73 | 25.1 | 0.24 | 0.97 | | 33.3 | | 0.16 | 0.48 |
| **4** | 25.3 | 0.11 | 0.45 | 26.1 | 0.17 | 0.67 | | 33.3 | | 0.16 | 0.49 |
| **5** | 26.1 | 0.20 | 0.75 | 27.0 | 0.28 | 1.04 | | 33.2 | | 0.22 | 0.68 |
| **6** | 27.3 | 0.16 | 0.58 | 28.2 | 0.21 | 0.73 | | 33.4 | | 0.18 | 0.53 |
| **7** | 28.2 | 0.15 | 0.52 | 29.1 | 0.21 | 0.73 | | 33.5 | | 0.19 | 0.56 |
| **8** | 29.2 | 0.15 | 0.5 | 30.1 | 0.14 | 0.47 | | 33.4 | | 0.21 | 0.64 |
| **9** | 30.2 | 0.11 | 0.37 | 31.0 | 0.15 | 0.49 | | 33.6 | | 0.18 | 0.53 |
| **10** | 31.0 | 0.22 | 0.71 | 31.9 | 0.26 | 0.80 | | 33.6 | | 0.18 | 0.53 |
| **11** | 31.9 | 0.15 | 0.45 | 33.0 | 0.20 | 0.61 | | 33.6 | | 0.14 | 0.43 |
| **12** | 32.7 | 0.16 | 0.49 | 33.8 | 0.20 | 0.58 | | 33.7 | | 0.16 | 0.47 |
| **R^2^** | **0.9987** | |  | **0.999** | | |  | |  |  |  |
| **13-NNW** | 0 |  |  | 0 |  |  | | 33.7 | | 0.25 | 0.76 |

ORF1a: Open Reading Frame 1a, E: Envelope, RNAseP: Ribonuclease P, C**_T_**: Cycle threshold, Avg: Average, SD: Standard Deviation, CV: Coefficient of Variation, NNW: Negative Nasal Wash, R^2^: r squared

**Table S5**: ORF1a, E and RNAse P values obtained for each of the 20 replicates using cobas® 6800 (Covid only) at Emory Molecular Laboratory (submitted as Table S5. csv file).

**Table S6**: Average ORF1a and E gene C_T_ values obtained for 20 replicates from cobas® SARS-CoV-2 & Influenza A/B Assay by Quest Diagnostics. R^2^ was calculated in excel with dilution number on y-axis and C_T_ on x-axis, in a scatterplot.

|  | **Roche Cobas ORF1a** | | | **Roche Cobas E** | | |
| --- | --- | --- | --- | --- | --- | --- |
| **Dilution** | **ORF1a C_T_ Avg** | **C_T_ SD** | **CV** | **E C_T_ Avg** | **C_T_ SD** | **CV** |
| **1** | 23.4 | 0.3 | 1.27 | 24.1 | 0.31 | 1.30 |
| **2** | 24.4 | 0.2 | 0.82 | 25.3 | 0.21 | 0.84 |
| **3** | 25.6 | 0.36 | 1.43 | 26.4 | 0.37 | 1.41 |
| **4** | 26.3 | 0.37 | 1.40 | 27.2 | 0.41 | 1.50 |
| **5** | 27.6 | 0.57 | 2.06 | 28.4 | 0.60 | 2.11 |
| **6** | 28.8 | 0.24 | 0.84 | 29.8 | 0.25 | 0.86 |
| **7** | 29.9 | 0.44 | 1.46 | 30.9 | 0.48 | 1.55 |
| **8** | 30.6 | 0.56 | 1.82 | 31.7 | 0.62 | 1.95 |
| **9** | 31.3 | 0.33 | 1.05 | 32.4 | 0.39 | 1.21 |
| **10** | 32.0 | 0.26 | 0.80 | 33.3 | 0.30 | 0.89 |
| **11** | 32.6 | 0.33 | 1.01 | 34.2 | 0.28 | 0.82 |
| **12** | 33.1 | 0.16 | 0.48 | 34.8 | 0.39 | 1.11 |
| **R^2^** | **0.9853** | |  | **0.9899** | |  |
| **13-NNW** | 0 |  |  | 0 |  |  |

ORF1a: Open Reading Frame 1a, E: Envelope, C**_T_**: Cycle threshold, Avg: Average, SD: Standard Deviation, CV: Coefficient of Variation, NNW: Negative Nasal Wash, R^2^: r squared

**Table S7**: ORF1a, and E gene C_T_ values for each of the 20 replicates obtained with cobas® SARS-CoV-2 & Influenza A/B Assay by Quest Diagnostics (submitted as Table S7. csv file).

**Table S8**: Average N1, N2 and RNASeP gene C_T_ values obtained for 20 replicates from BD MAX System by TriCore Reference Laboratories. R^2^ was calculated in excel with dilution number on y-axis and C_T_ on x-axis, in a scatterplot.

|  | **BD Max N1** | | | **BD Max N2** | | | **BD Max RNaseP** | | |
| --- | --- | --- | --- | --- | --- | --- | --- | --- | --- |
| **Dilution** | **N1 C_T_ Avg** | **C_T_ SD** | **CV** | **N2 C_T_ Avg** | **C_T_ SD** | **CV** | **RNaseP C_T_ Avg** | **C_T_ SD** | **CV** |
| **1** | 21.4 | 0.21 | 1.0 | 22.0 | 0.24 | 1.1 | 25.7 | 0.35 | 1.4 |
| **2** | 21.3 | 0.13 | 0.6 | 21.7 | 0.26 | 1.2 | 25.9 | 0.19 | 0.7 |
| **3** | 23.3 | 0.16 | 0.7 | 24.1 | 0.27 | 1.1 | 25.8 | 0.26 | 1.0 |
| **4** | 23.2 | 0.23 | 1.0 | 23.9 | 0.30 | 1.3 | 25.7 | 0.34 | 1.3 |
| **5** | 24.6 | 0.22 | 0.9 | 25.3 | 0.15 | 0.6 | 26.1 | 0.17 | 0.7 |
| **6** | 25.8 | 0.10 | 0.4 | 26.6 | 0.20 | 0.7 | 26.5 | 0.17 | 0.6 |
| **7** | 26.0 | 0.27 | 1.0 | 26.7 | 0.46 | 1.7 | 26.0 | 0.29 | 1.1 |
| **8** | 27.2 | 0.16 | 0.6 | 28.1 | 0.23 | 0.8 | 26.4 | 0.38 | 1.4 |
| **9** | 29.5 | 0.37 | 1.2 | 30.2 | 0.41 | 1.4 | 26.5 | 0.28 | 1.1 |
| **10** | 29.5 | 0.35 | 1.2 | 30.4 | 0.33 | 1.1 | 26.4 | 0.40 | 1.5 |
| **11** | 30.3 | 0.28 | 0.9 | 31.3 | 0.35 | 1.1 | 25.8 | 0.45 | 1.7 |
| **12** | 31.1 | 2.02 | 6.5 | 32.1 | 0.42 | 1.1 | 26.0 | 0.24 | 0.9 |
| **R^2^** | **0.9462** | |  | **0.9846** | |  |  |  |  |
| **13-NNW** | 0 |  |  | 0 |  |  | 33.7 | 0.25 | 0.76 |

N1: N1 region of the nucleocapsid gene, N2: N2 region of the nucleocapsid gene, RNAseP: Ribonuclease P, C**_T_**: Cycle threshold, Avg: Average, SD: Standard Deviation, CV: Coefficient of Variation, , R^2^: r squared, NNW: Negative Nasal Wash, BD: Becton Dickinson.

**Table S9:** N1 and N2 C_T_ values for each of the 20 replicates obtained BD MAX System by TriCore Reference Laboratories. (Submitted as Table S9. csv file).

**Table S10:** Average E, N2 and RdRp gene values for 20 replicates using Xpert Xpress CoV-2 plus by Cepheid at ELIAD. R^2^ was calculated in excel with dilution number on y-axis and C_T_ on x-axis, in a scatterplot.

|  | **Cepheid E** | | | **Cepheid N2** | | | **Cepheid RdRp** | | |
| --- | --- | --- | --- | --- | --- | --- | --- | --- | --- |
| **Dilution** | **E C_T_ Avg** | **C_T_ SD** | **CV** | **N2 C_T_ Avg** | **C_T_ SD** | **CV** | **RNaseP C_T_ Avg** | **C_T_ SD** | **CV** |
| **1** | 21.1 | 0.18 | 0.8 | 24.9 | 0.21 | 0.86 | 23.4 | 0.18 | 0.8 |
| **2** | 22.3 | 0.21 | 1.0 | 26.0 | 0.24 | 0.94 | 24.7 | 0.23 | 1.0 |
| **3** | 23.3 | 0.19 | 0.8 | 27.0 | 0.25 | 0.91 | 25.7 | 0.25 | 1.0 |
| **4** | 24.4 | 0.23 | 0.9 | 28.2 | 0.23 | 0.82 | 26.9 | 0.32 | 1.2 |
| **5** | 25.4 | 0.14 | 0.5 | 29.1 | 0.20 | 0.69 | 27.8 | 0.21 | 0.8 |
| **6** | 26.6 | 0.44 | 1.7 | 30.4 | 0.47 | 1.54 | 29.1 | 0.47 | 1.6 |
| **7** | 27.6 | 0.23 | 0.8 | 31.3 | 0.25 | 0.8 | 30.1 | 0.22 | 0.7 |
| **8** | 28.9 | 0.83 | 2.9 | 32.5 | 0.50 | 1.54 | 31.4 | 0.74 | 2.4 |
| **9** | 29.9 | 0.26 | 0.9 | 33.7 | 0.25 | 0.73 | 32.5 | 0.25 | 0.8 |
| **10** | 30.8 | 0.29 | 1.0 | 34.4 | 0.32 | 0.94 | 33.3 | 0.28 | 0.8 |
| **11** | 31.9 | 0.35 | 1.1 | 35.6 | 0.29 | 0.82 | 34.5 | 0.46 | 1.3 |
| **12** | 32.9 | 0.32 | 1.0 | 36.7 | 0.34 | 0.93 | 35.5 | 0.32 | 0.9 |
| **R^2^** | **0.9995** | |  | **0.9993** | |  | **0.9992** | |  |
| **13-NNW** | 0 |  |  | 0 |  |  | 33.7 | 0.25 | 0.76 |

E: Envelope, N2: N2 region of the nucleocapsid gene, RdRp- RNA dependent RNA Polymerase, C**_T_**: Cycle threshold, Avg: Average, SD: Standard Deviation, CV: Coefficient of Variation, NNW: Negative Nasal Wash, R^2^: r squared

**Table S11:** E, N2 and RdRp C_T_ values for each of the 20 replicates obtained from Cepheid by ELIAD. (Submitted as Table S11. csv file).

**Table S12:** Average RTPCR values for 20 replicates using N2 and SC2 CDC primers. R^2^ was calculated in excel with dilution number on y-axis and C_T_ on x-axis, in a scatterplot.

|  | **N2 (CDC) Primers** | | | **SC2 (CDC) Primers** | | |
| --- | --- | --- | --- | --- | --- | --- |
| **Dilution** | **N2 C_T_ Avg** | **C_T_ SD** | **CV** | **SC2 C_T_ Avg** | **C_T_ SD** | **CV** |
| 1 | 22.2 | 0.12 | 0.53 | 23.0 | 0.06 | 0.28 |
| 2 | 23.5 | 0.09 | 0.39 | 24.3 | 0.12 | 0.48 |
| 3 | 24.6 | 0.11 | 0.45 | 25.4 | 0.15 | 0.59 |
| 4 | 25.7 | 0.15 | 0.60 | 26.4 | 0.13 | 0.51 |
| 5 | 26.1 | 0.07 | 0.27 | 26.8 | 0.08 | 0.28 |
| 6 | 27.1 | 0.20 | 0.73 | 27.7 | 0.19 | 0.68 |
| 7 | 28.8 | 0.13 | 0.45 | 29.0 | 0.08 | 0.28 |
| 8 | 29.4 | 0.11 | 0.39 | 29.6 | 0.12 | 0.42 |
| 9 | 30.5 | 0.13 | 0.43 | 30.3 | 0.16 | 0.53 |
| 10 | 31.7 | 0.20 | 0.65 | 31.2 | 0.14 | 0.45 |
| 11 | 32.7 | 0.30 | 0.93 | 31.8 | 0.21 | 0.65 |
| 12 | 33.9 | 0.36 | 1.07 | 32.5 | 0.21 | 0.64 |
| **R2** | **0.9967** | |  | **0.9931** | |  |
| 13- NNW | 0.00 |  |  | 0.0 |  |  |

N2: N2 region of the nucleocapsid gene, SC2: Influenza SARS-CoV-2 Multiplex assay, CDC: Center for Disease Control, C**_T_**: Cycle threshold, Avg: Average, SD: Standard Deviation, CV: Coefficient of Variation, NNW: Negative Nasal Wash, R^2^: r squared

**Table S13:** N2 and SC2 individual C_T_ values obtained for 20 replicates using the N2 and SC2 primer probe sets. (Submitted as Table S13. csv file).

Individual C_T_ values submitted as supplementary data files were used in Passing-Bablok and Deming Regression analysis ^1,2^.

As we saw, Tables S4 to S13 contained both the individual and the average C_T_ values across different labs using various comparator platforms. The following section details the calculations used for the 30 C_T_ equivalents for several assays.

**Assessing Passing-Bablok Regression Fit on Roche cobas® E C_T_ vs. Natural-Log Transformed ddPCR After Removing Outliers**

After the circled outliers were removed, Passing-Bablok regression shows a slope of 1.07 and an intercept of 8.55 as seen in Figure S1 below.

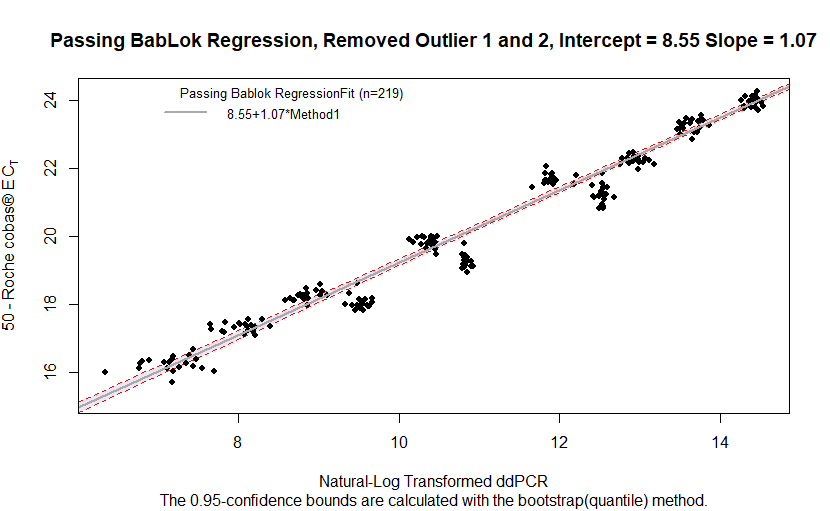

**Figure S1.** Passing-Bablok Regression with log-transformed ddPCR GE/ml concentration on the x-axis and 50 – Roche **cobas®** E C_T_ on the y-axis after outliers have been removed.

**Comparing the Performance of Deming Regression on dilutions analyzed by BD Max instrument at TriCore to the Performance on dilutions analyzed using** cobas® **SARS-CoV-2 Test at Emory Molecular Labs.**

Comparing the Passing-Bablok regression plots for N1 and N2 C_T_ obtained using BD Max at TriCore, we see that the BD N1 regression model left three outlier samples to be removed while the BD N2 regression model left only one outlier sample to be removed.

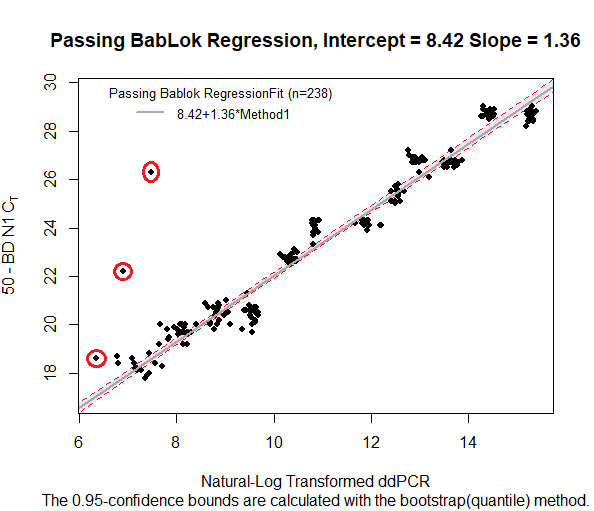

**Figure S2.** Passing-Bablok Regression with log-transformed ddPCR GE/ml on the x-axis and 50 – BD N1 C_T_ on the y-axis. Outliers to be removed are denoted with red circles.

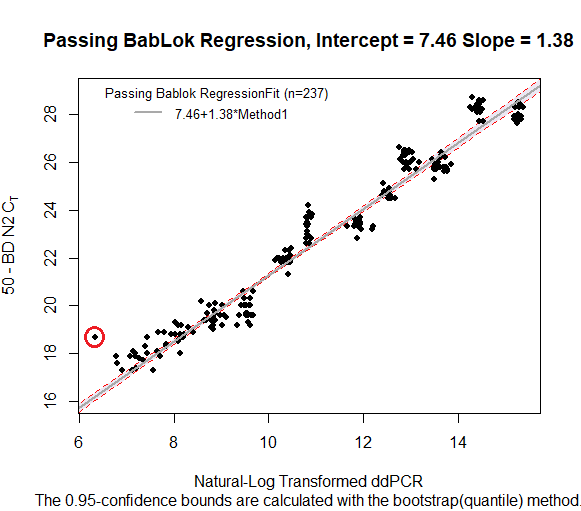

**Figure S3.** Passing-Bablok Regression with log-transformed ddPCR GE/ml on the x-axis and 50 – BD N2 C_T_ on the y-axis. Outliers to be removed are denoted with red circles.

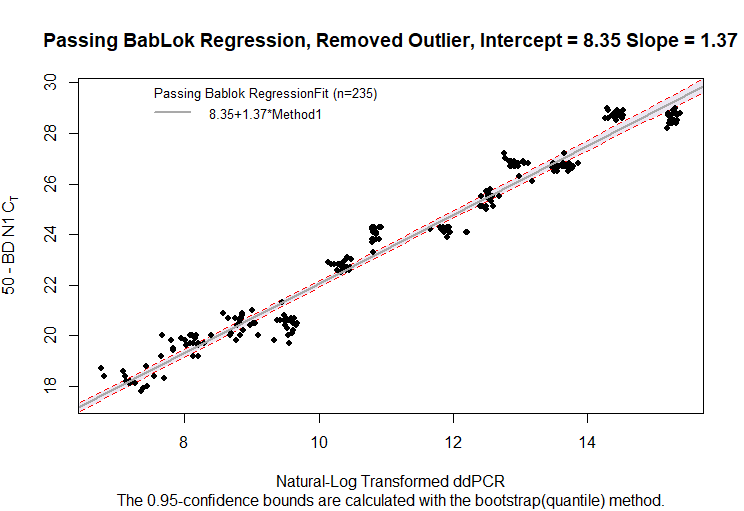

**Figure S4.** Passing-Bablok Regression with log-transformed ddPCR GE/ml on the x-axis and 50 – BD N1 C_T_ on the y-axis after outliers were removed.

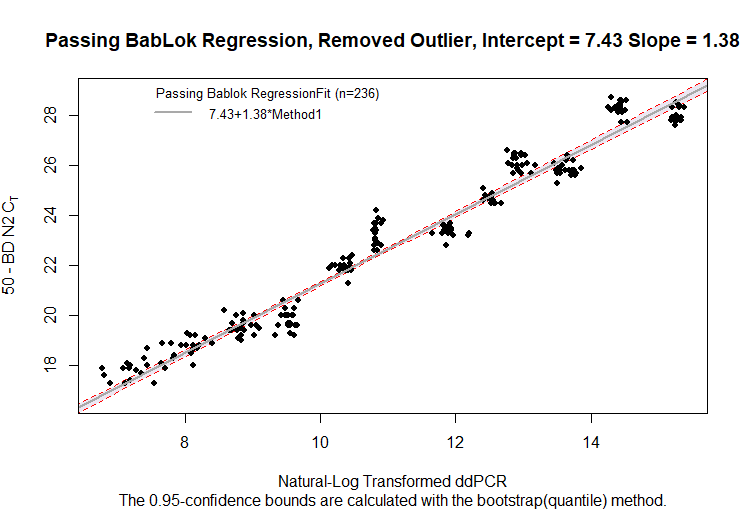

**Figure S5.** Passing-Bablok Regression with log-transformed ddPCR GE/ml on the x-axis and 50 – BD N2 C_T_ on the y-axis after the outlier was removed.

From the table below, we see that the delta parameter we should use in the Deming regression model for BD Max N1 is 9.406 while for BD Max N2 is 19.246. Dilution 7 produced noticeably high variance ratios for both BD Max N1 (46.961) and BD Max N2 (139.215) and seemed to have significant influence on the delta parameter we chose to use for each model.

**Table S14.** Calculations of variance and standard deviation ratios between the variance of the log-transformed ddPCR GE/ml and the transformed BD N1 and BD N2 C_T_ values along with their respective averages.

| Dilution | BD Max N1 Delta: Var(Y1)/Var(X) | BD Max N2 Delta: Var(Y2)/Var(X) | BD Max N1 CV(Y1)/CV(X) | BD Max N2 CV(Y2)/CV(X) |
| --- | --- | --- | --- | --- |
| 1 | 15.192 | 21.348 | 3.898 | 4.620 |
| 2 | 2.694 | 11.593 | 1.641 | 3.405 |
| 3 | 1.901 | 5.435 | 1.379 | 2.331 |
| 4 | 4.582 | 7.698 | 2.140 | 2.775 |
| 5 | 12.418 | 5.966 | 3.524 | 2.443 |
| 6 | 0.626 | 2.656 | 0.791 | 1.630 |
| 7 | 46.961 | 139.215 | 6.853 | 11.799 |
| 8 | 2.664 | 5.952 | 1.632 | 2.440 |
| 9 | 16.25 | 20.202 | 4.031 | 4.495 |
| 10 | 6.419 | 5.766 | 2.534 | 2.401 |
| 11 | 1.857 | 2.958 | 1.363 | 1.720 |
| 12 | 1.305 | 2.166 | 1.142 | 1.472 |
| Average | 9.406 | 19.246 | 2.577 | 3.461 |

Deming regression on the two BD instruments produced nearly identical slopes (1.37 for BD N1 and 1.38 for BD N2), while the intercepts were slightly different (8.41 for BD N1 and 7.55 for BD N2).

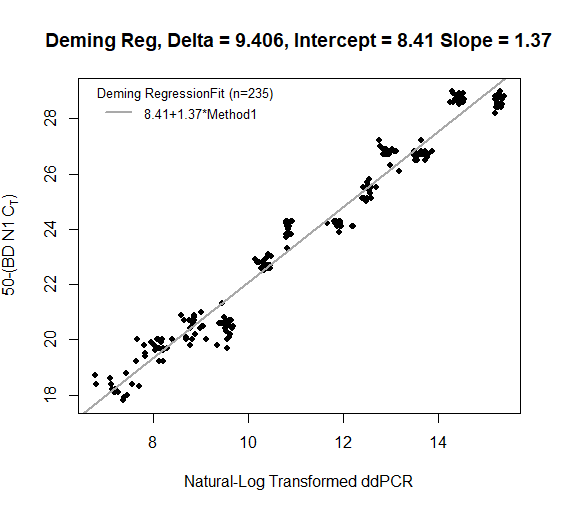

**Figure S6.** Deming Regression of Data without outliers with log-transformed ddPCR GE/ml on the x-axis and 50 – BD Max N1 C_T_ on the y-axis, using the computed average ratio of the variances as the delta.

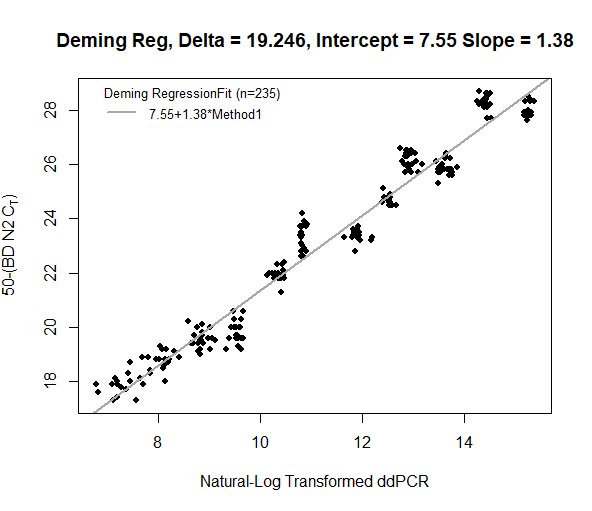

**Figure S7.** Deming Regression of Data without outliers with log-transformed ddPCR GE/ml on the x-axis and 50 – BD N2 C_T_ on the y-axis, using the computed average ratio of the variances as our delta.

After performing the appropriate calculations, we find that the Roche cobas® E2 30 C_T_ equivalent for BD Max N1 is 26.83 and for BD Max N2 is 27.54.

BD Max N2 Example Calculation

(50 – X) = 7.55 + 1.38*ln(49447)

X = 42.45 – 1.38*ln(49447)

X = 27.54 C_T_

**Comparing the Performance of Deming Regression on dilutions analyzed by Emory CDC instruments to the Performance on dilutions analyzed using** cobas® **SARS-CoV-2 Test at Emory Molecular Labs.**

After plotting the results of Passing-Bablok regression performed on the Emory CDC N2 and CDC SC2 instruments, we see that no outliers were present in the plot using CDC N2, but we do see a cluster of outlier samples present in the model using CDC SC2.

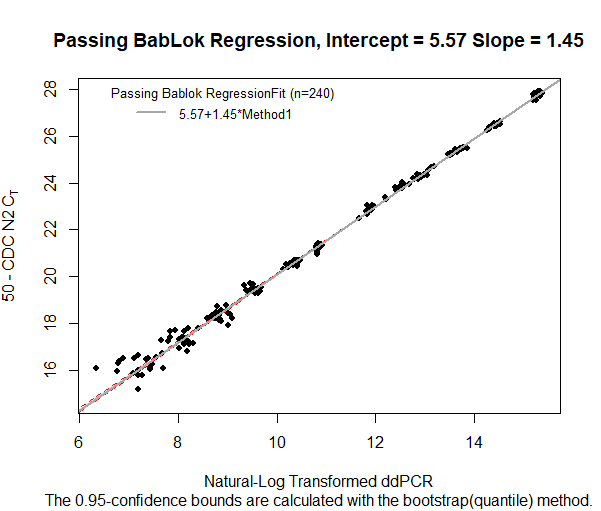

**Figure S8.** Passing-Bablok Regression with log-transformed ddPCR GE/ml on the x-axis and 50 – CDC N2 C_T_ on the y-axis.

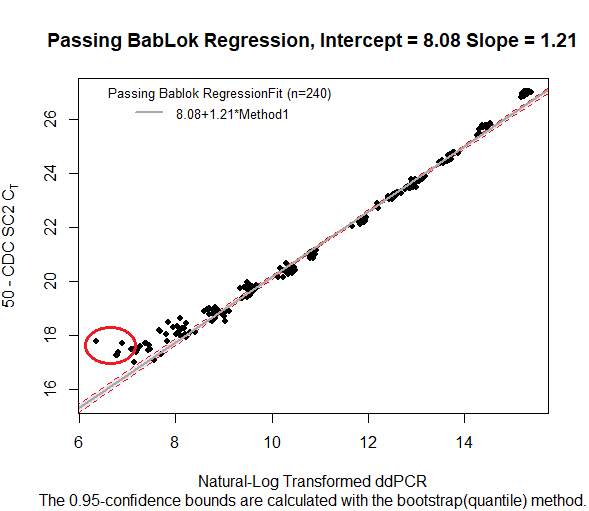

**Figure S9.** Passing-Bablok Regression with log-transformed ddPCR GE/ml on the x-axis and 50 – CDC SC2 C_T_ on the y-axis. Outliers to be removed are denoted with red circles.

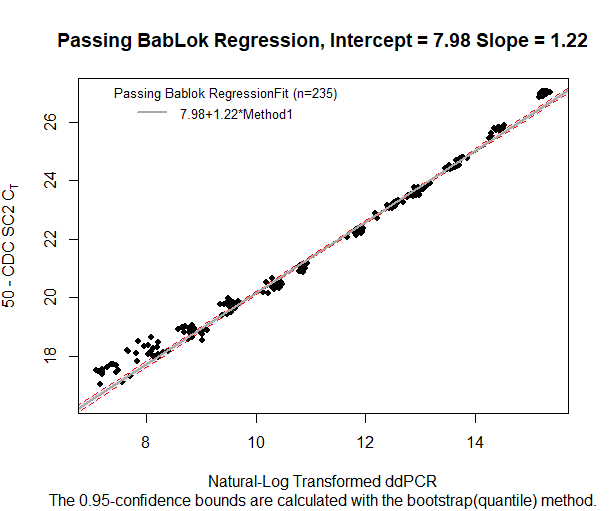

**Figure S10.** Passing-Bablok Regression with log-transformed ddPCR GE/ml on the x-axis and 50 – CDC SC2 C_T_ on the y-axis after outliers were removed.

From the following table below, we find that the delta parameter we should use in the Deming regression model for CDC N2 is 3.11 and for CDC SC2 is 1.97. Dilution 7 again provided slightly more extreme variance ratios for both Emory instruments, but they were not as noteworthy as in the previous tables in this analysis.

**Table S15.** Calculations of variance and standard deviation ratios between the variance of the log-transformed ddPCR GE/ml and the transformed CDC N2 and CDC SC2 C_T_ values along with their respective averages.

| Dilution | Var(X): Var(Log(ddPCR)) | | CDC N2 Delta: Var(Y1)/Var(X) | CDC SC2 Delta: Var(Y2)/Var(X) | | CV(Y1)/CV(X) | CV(Y2)/CV(X) |
| --- | --- | --- | --- | --- | --- | --- | --- |
| 1 | | 0 | 4.93 | 1.46 | 2.22 | | 1.21 |
| 2 | | 0.01 | 1.36 | 2.22 | 1.17 | | 1.49 |
| 3 | | 0.01 | 0.93 | 1.75 | 0.97 | | 1.32 |
| 4 | | 0.01 | 2.07 | 1.57 | 1.44 | | 1.25 |
| 5 | | 0 | 1.28 | 1.49 | 1.13 | | 1.22 |
| 6 | | 0.01 | 2.73 | 2.49 | 1.65 | | 1.58 |
| 7 | | 0 | 11.37 | 4.6 | 3.37 | | 2.14 |
| 8 | | 0.01 | 1.4 | 1.68 | 1.18 | | 1.30 |
| 9 | | 0.01 | 2.06 | 3.09 | 1.44 | | 1.76 |
| 10 | | 0.02 | 2.21 | 1.04 | 1.49 | | 1.02 |
| 11 | | 0.04 | 2.25 | 1.04 | 1.50 | | 1.02 |
| 12 | | 0.03 | 4.72 | 1.21 | 2.17 | | 1.10 |
| Average | | 0.01 | 3.11 | 1.97 | 1.64 | | 1.37 |

The Deming regressions for both Emory instruments produced similar slopes (1.44 for CDC N2 and 1.20 for CDC SC2) but quite different intercepts (5.68 for CDC N2 and 8.28 for CDC SC2).

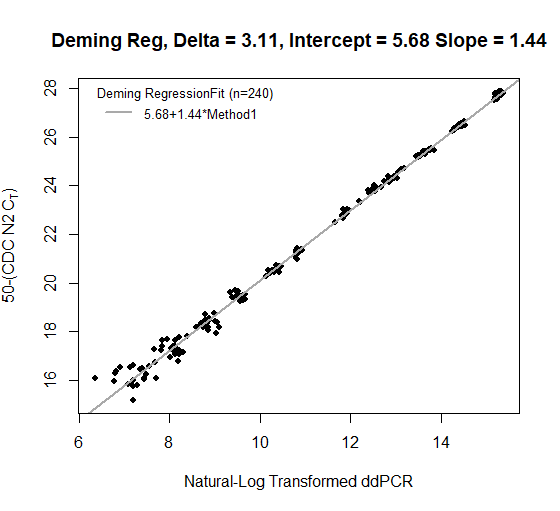

**Figure S11.** Deming Regression of Data without outliers with log-transformed ddPCR GE/ml on the x-axis and 50 – CDC N2 C_T_ on the y-axis, using the computed average ratio of the variances as our delta.

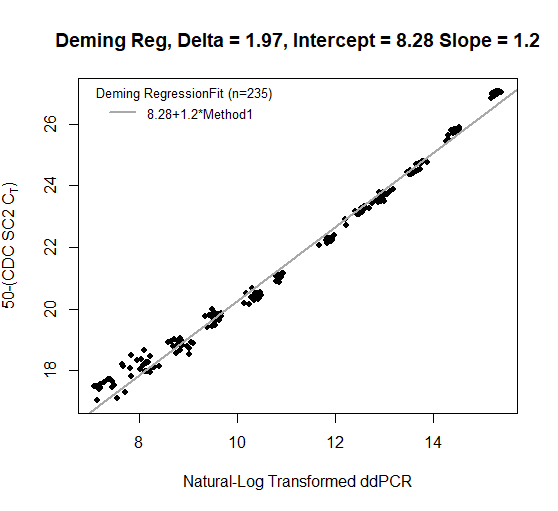

**Figure S12.** Deming Regression of Data without outliers with log-transformed ddPCR GE/ml on the x-axis and 50 – CDC SC2 C_T_ on the y-axis, using the computed average ratio of the variances as our delta.

Using the same formula to find the calibrated C_T_ values for BD N1 and BD N2, we see that the Roche E2 30 C_T_ equivalent for CDC N2 is 28.72 and for CDC SC2 is 28.77. Comparing these results to the calibrated C_T_ values for BD N1 and BD N2, we see that the Emory instruments produced calibrated C_T_ s closer to 30 than the TriCore instruments did, suggesting potentially stronger performance.

**Suppplement References**

1.	Payne, R. B. Method Comparison: Evaluation of Least Squares, Deming and Passing/Bablok Regression Procedures Using Computer Simulation. *Ann. Clin. Biochem.* **34**, 319–320 (1997).

2.	Redmond, T., Russell, R. A., Anderson, R. S. & Garway-Heath, D. F. Author Response: Passing–Bablok Regression Is Inappropriate for Assessing Association Between Structure and Function. *Invest. Ophthalmol. Vis. Sci.* **54**, 5850–5851 (2013).
